# Molecular PD-L1 PET/CT Imaging with ^89^Zr-atezolizumab to Monitor Immune Responses in metastatic Triple Negative Breast Cancer: study protocol for a phase II diagnostic imaging trial

**DOI:** 10.1101/2023.03.13.23287203

**Authors:** Renske Altena, Thuy A. Tran, Una Kjällquist, Mattias Karlsson, Jonathan Siikanen, Emma Jussing, Antonios Tzortzakakis, Olof Jonmarker, Rimma Axelsson, Jonas Bergh

**Author notes:** Correspondence: Renske Altena MD PhD, Medical Unit Breast, Endocrine tumors and Sarcoma, Karolinska University Hospital Solna, Sweden.

## Abstract

**Background:** Immune checkpoint inhibitors (Programmed Death Ligand 1 [PD-L1] and Programmed Death 1 [PD-1] antibodies) are approved for the first line treatment of metastatic and irresectable triple negative breast cancer (mTNBC) with immunohistochemical (IHC) PD-L1 positivity on a tumor biopsy, and combined with a chemotherapy backbone. Intra- and interlesional as well as spatial heterogeneity in PD-L1 expression as well the invasive nature of tumor biopsies poses disadvantages of this method of selecting patients for the addition of PD-L1/PD-1 antibodies. Our study hypothesis is that whole body Positron Emission Tomography with computed tomography (PET/CT) imaging with a PD-L1 specific radiotracer could be a more reliable biomarker approach for the selection of patients with mTNBC who benefit from the addition from PD-L1/PD-1 antibodies.

**Methods:** In the phase II “Molecular PD-L1 PET/CT Imaging with ^89^Zr-atezolizumab to Monitor Immune Responses in metastatic Triple Negative Breast Cancer” (MIMIR-mTNBC) trial, treatment enrichment for checkpoint inhibitors is done by a baseline ^89^Zr-atezolizumab PET/CT followed by a tumor biopsy to determine PD-L1 by IHC. Patients with a PD-L1 positive tumor on biopsy and/or PET are treated with chemotherapy + atezolizumab. The primary aim of the study is to assess the level of statistical agreement between the reference standard (PD-L1 IHC) compared to the experimental method (PD-L1 PET) by means of Cohen’s kappa coefficient. The trial is registered at Clinicaltrials.org with number NCT05742269.

**Discussion:** The results of the MIMIR-mTNBC trial are expected to reveal the possibility of PD-L1 PET with ^89^Zr-atezolizumab to complement or replace the assessment of PD-L1 status by means of IHC on a tumor biopsy, and as a possible method to select patients with mTNBC that benefit from the addition of atezolizumab to a chemotherapy backbone. This non-invasive tool may provide a method that reveals relevant information about heterogeneity in the expression of the target of treatment.

**Trial registration:** NCT05742269, Trial registered on February 23^rd^, 2023, at clinicaltrials.gov

## Introduction

The current standard of care for triple negative breast cancer includes the combination of immune checkpoint inhibitors (ICI) with a chemotherapy backbone, both in the neo-adjuvant and metastatic setting. For patients with metastatic or irresectable triple negative breast cancer (mTNBC) atezolizumab (Programmed Death Ligand 1 [PD-L1]) and pembrolizumab (anti Programmed Death [PD-1]) are approved for patients with immunohistochemical (IHC) PD-L1 expression in a tumor biopsy according to specific criteria with a companion diagnostic IHC antibody, based on data from the IMpassion130 and Keynote355 trials respectively (1-4).

In the IMpassion130 study, patients with mTNBC were treated with nab-paclitaxel atezolizumab or placebo. A statistically significant improvement in progression free survival (PFS) and a clinically meaningful improvement in overall survival (OS) for patients whose tumor had over 1% PD-L1 expression on tumor infiltrating lymphocytes was found, with a numeric improvement in median PFS of 2.5 months (7.5 months and 5.0 months, respectively with a hazard ratio [HR] of 0.62; 95% confidence interval [CI] 0.49-0.78; P<0.001) and a median OS of almost 10 months (25.0 months and 15.5 months, respectively, with a HR of 0.62; 95% CI, 0.45-0.86) (1,2). At the final OS update, this benefit in overall survival landed at 7.5 months in the PD-L1 positive subgroup (HR, 0.67; 95% CI, 0.53-0.86) (2).

In the Keynote355 study, patients with a Combined Positive Score (CPS with the pharmDx test) of 10 or more treated with pembrolizumab plus chemotherapy (either nab-paclitaxel; paclitaxel; or gemcitabine plus carboplatin) had an improvement in median PFS of little over 4 months (9.7 months and 5.6 months respectively, HR for progression or death, 0.65, 95% CI 0.49–0.86) (3), and an increase in OS for patients in the CPS-10 subgroup was from 16.1 months in the placebo–chemotherapy group to 23.0 months in the pembrolizumab–chemotherapy group (hazard ratio for death, 0.73; 95% CI 0.55-0.95) (4).

There is an urgent clinical need to establish reliable and reproducible therapy-predictive biomarkers for ICI in solid tumors, in order to optimize the risk-benefit balance on an individual and societal level. Up to now, for mTNBC the only approved biomarker for the addition of ICI to a chemotherapy backbone is PD-L1 expression on a tumor biopsy according to pre-defined criteria as mentioned above [5]. However, this method also has a suboptimal performance, as concluded in a systematic review and meta-analysis across all solid tumors [6]. For TNBC, several alternative candidate biomarkers have been proposed but have yet to be validated thoroughly enough to be implemented in clinical practice and replace PD-L1 IHC on a tumor biopsy [7]. It has been unequivocally described that PD-L1 expression can change over time in primary and metastatic breast cancer, as well as have varying levels of expression in metastases in different organs [8], which limits even further the therapy-predictive role of PD-L1 IHC for the benefit of ICIs for patients with TNBC. Interestingly, PD-L1 status in the primary tumor has no therapy-predictive role for the addition of pembrolizumab to a chemotherapy backbone in the neoadjuvant treatment setting, as was observed in the Keynote522 trial [9].

Positron Emission Tomography (PET) imaging with receptor-specific tracers can provide, in a non-invasive manner, information on whether the target for treatment exists. At the same time, PET enables quantification of the target in the whole body in real-time and with a high degree of accuracy. Several radiotracers representing treatment targets in breast cancer management have been evaluated, e.g. tracers visualizing the Estrogen Receptor (ER) and Human Epidermal growth factor Receptor 2 (HER2). 18F-FES, representing ER-positive lesions, is a currently FDA approved tracer that can be used in combination with a tumor biopsy to determine ER status in the whole body (10).

A few radiotracers representing targets of ICIs, or their downstream intracellular effects, have been studied [11]. The first study published is a first-in-human study with ^89^Zr-atezolizumab in 25 patients (NCT02453984), including four patients with mTNBC [12]. This study showed that imaging with the tracer was feasible and safe. The results indicated clear heterogeneity of expression within and between lesions and a promising role as a predictive marker for response to treatment with PD-L1 antibodies. For the patients with mTNBC in this study, SUVmax was clearly over the SUVmean in the blood pool measured over the aorta, with varying levels of SUVmax in different organs. In this study, higher SUVmax was related to a better antitumor response, according to RECIST.

Here we report the study protocol of the Molecular Imaging to Monitor Immune Responses in mTNBC (MIMIR-mTNBC; EU CT 2022-500808-21-00, NCT05742269) trial. In this phase II diagnostic imaging study, we aim to evaluate the role of ^89^Zr-atezolizumab PET/CT as a way to refine the group of patients with mTNBC that benefit from the addition of an ICIs to a chemotherapy backbone. The primary purpose of the trial is to establish the level of statistical agreement between PD-L1 status by the current reference standard with IHC versus PET with ^89^Zr-atezolizumab. Our hypothesis is that Positron Emission Tomography combined with Computed Tomography (PET/CT) imaging with a contemporary radiotracer (^89^Zr-atezolizumab), visualizing PD-L1 expression in the whole body, could be a better predictive biomarker to select which patients benefit from ICI.

## Materials and methods

To test this hypothesis, we are performing a phase II PD-L1 PET/CT imaging study with ^89^Zr-atezolizumab in patients with mTNBC. The study design is summarized as SPIRIT schedule of enrolment, interventions and assessments in Figure 1. A graphical representation of the study design is presented in Figure 2.

**Figure 1.**
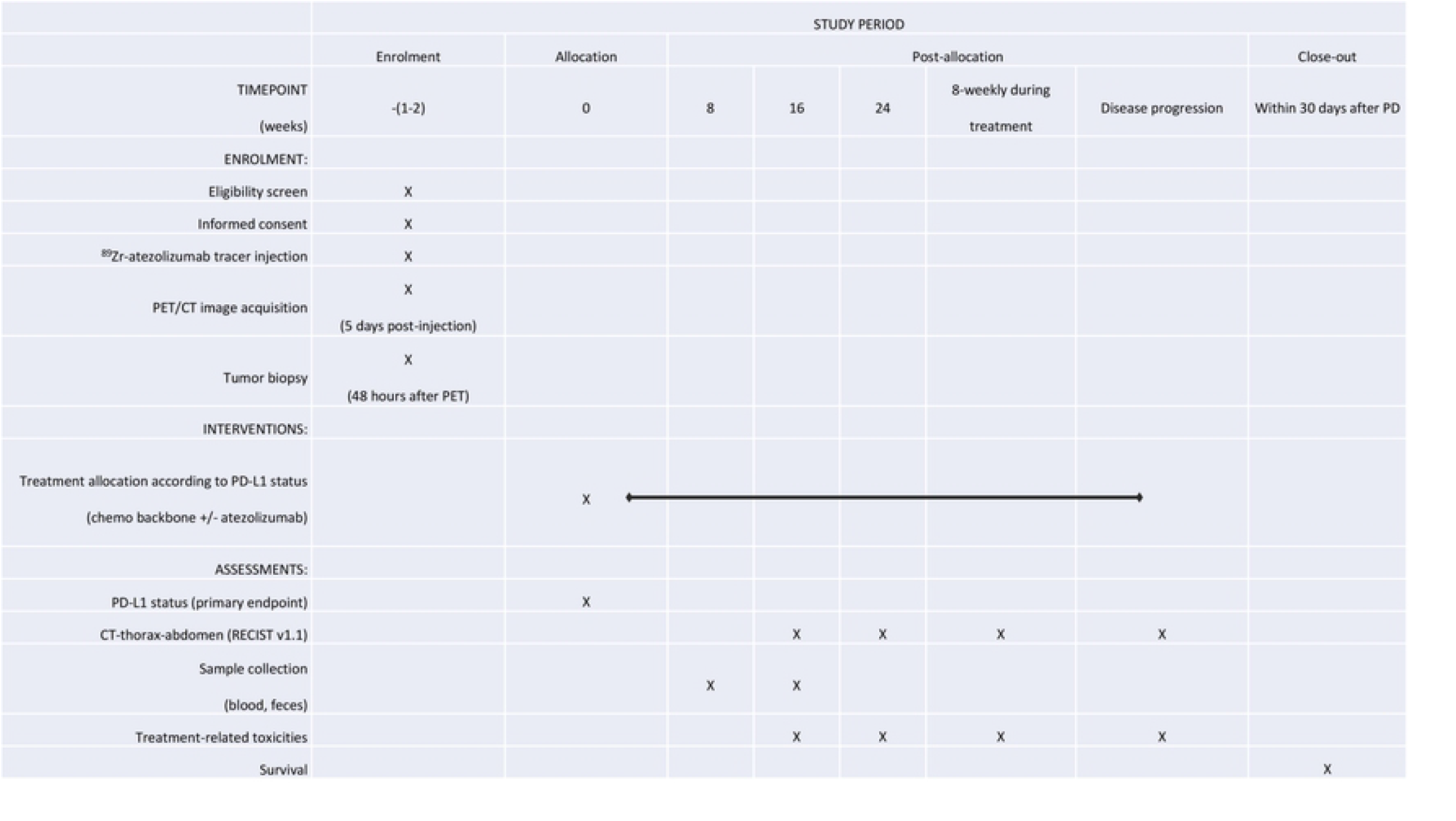
SPIRIT schedule of enrolment, interventions and assessments in the MIMIR-mTNBC trial. *Abbreviations: programmed death ligand 1 (PD-L1), Positron Emission Tomography with Computed Tomography (PET/CT), progressive disease (PD)*

**Figure 2:**
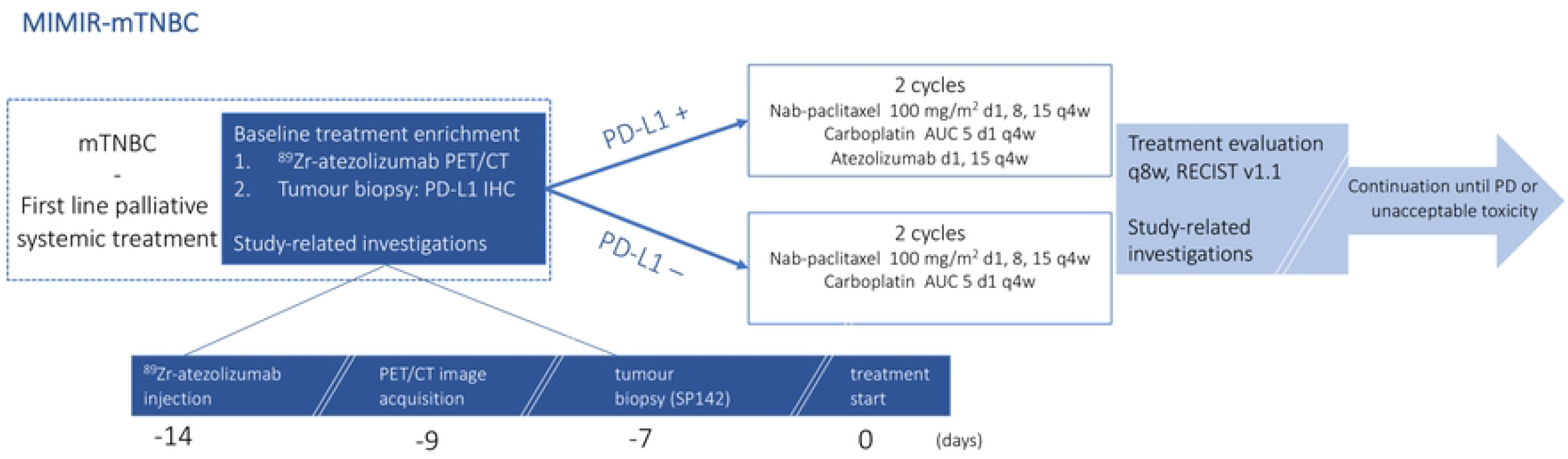
Study design of the MIMIR-mTNBC trial. *Abbreviations: metastatic triple negative breast cancer (mTNBC), programmed death ligand 1 (PD-L1), Positron Emission Tomography with Computed Tomography (PET/CT), immunohistochemistry (IHC), area under the curve (AUC)*.

### Study setting

Our target population consists of patients with mTNBC scheduled for first line palliative systemic treatment with nab-paclitaxel and carboplatin. This chemotherapy combination is used to maximize the therapeutic potential of this first line systemic treatment line, extrapolating signals from early TNBC (13) and in the absence of signs that indicate augmented safety issues (14).

Patients will be recruited at Karolinska University Hospital in Stockholm, Sweden. Additional Swedish study sites are planned to be added. The study-specific investigations with ^89^Zr-atezolizumab PET/CT, the study biopsies and blood samples will be done at the respective sites. The ^89^Zr-atezolizumab radiotracer will be centrally produced at the Department of Radiopharmacy, Karolinska University hospital in Solna, and shipped by courier to the respective sites throughout the country. The PD-L1 IHC will be centrally reviewed by the study pathologist that is dedicated to this study and trained and certified by Ventana on PD-L1 status assessment using SP142 and IHC. The PET-images will be acquired according to a standardized protocol that will be the same throughout all the sites and reviewed by a nuclear medicine physician ^89^Zr-atezolizumab radiotracer.

### Study population

Patients with metastatic/irresectable triple negative breast cancer are candidates for inclusion in the MIMIR-mTNBC trial, according to the inclusion and exclusion criteria summarized in Table 1. In short, patients must be fit and willing to receive treatment with chemotherapy plus/minus atezolizumab and undergo a tumor biopsy and PET/CT. The tumor localizations have to be measurable according to RECIST v1.1 (14) and a non-skeletal lesion has to be accessible for ultrasound- or CT-directed biopsy. No contra-indications for PET/CT investigations or treatment with atezolizumab must be present. Informed consent is mandatory before randomization and will be obtained by a physician with the appropriate delegation.

**Table 1.** Inclusion and exclusion criteria in the MIMIR-mTNBC trial.

| Inclusion criteria | Exclusion criteria |
| --- | --- |
| • Age ≥ 18 years<br>• Diagnosed with mTNBC (tumor biology: ER<10%, PR<10%, HER2-)<br>• Measurable disease according to RECIST v1.1<br>• At least one metastatic lesion accessible for biopsy<br>• Deemed by treating physician as fit for systemic therapy according to study protocol<br>• ECOG performance score 0 or 1<br>• Adequate blood tests for bone marrow, renal and hepatic functions (Table 6)<br>• Able and willing to provide written informed consent | • Previous treatment with chemotherapy or targeted therapy for mTNBC. Radiation therapy and previous chemotherapy (including taxanes) in the context of curative therapy (if treatment was completed ≥12 months before randomization) is allowed.<br>• Contraindications for PET/CT as defined for clinical practice<br>• Other malignancy within the past five years<br>• For women in childbearing age: pregnancy or lactation, or inadequate contraception<br>• Uncontrolled hypertension, heart-, liver-, or kidney-diseases or other medical/psychiatric disorders<br>• History of autoimmune disease with the exception of controlled hypothyroidism, type 1diabetes mellitus or skin diseases.<br>• Treatment with systemic immunosuppressive drugs |

### Experimental investigations

Study-related assessments include imaging, laboratory tests (blood, feces and tumour samples) and clinical outcome assessments including treatment response and toxicities, as summarized in the SPIRIT chart in Figure 1.

#### 1. Imaging Assessments

The participants of the study will undergo one session of ^89^Zr-atezolizumab PET/CT (PD-L1 PET in the rest of this manuscript), which is performed within 14 days before initiation of the systemic oncological treatment. Response evaluation after 2 courses of systemic therapy will be performed according to clinical routine with CT of chest and abdomen.

^89^Zr-atezolizumab is administered as a single intravenous injection at the department of Nuclear Medicine. Pre-injection as well as one hour after injection, blood pressure, pulse and temperature are taken by a nurse experienced with the administration of radiotracers. The patient can go home afterwards and will be phoned by the study nurse 48-72 hours later to follow-up clinical status.

At 5 days post-injection of ^89^Zr-atezolizumab, a whole-body PET will be acquired. Imaging will be performed using the same protocol on either a Siemens Biograph mCT PET/CT scanner (Siemens, Erlangen, Germany) or GE Discovery MI, (GE, Milwaukee, WI, USA) at participating sites (according to 2.4). The images acquired using PD-L1 PET will be analyzed with image processing software available in the clinic (Syngo.Via, Siemens Healthineers and Software from Hermes Medical Solution AB, Sweden), according to the following steps:

1. At the imaging day, a first examination will be done to assess whether ^89^Zr-atezolizumab PD-L1 positive lesions are found. This will be communicated with the study interventional radiologist who then decides the optimal location to obtain the study specific tumor biopsy.
2. The maximum, peak, and average standardized uptake value (SUVmax, SUV peak, and SUVmean), and Total PD-L1 Positive Volume) will be estimated in the whole-body images acquired and will be analysed after all patients have been included and results for the final analysis are prepared.

#### 2. Biological material: tumor, blood and feces samples

Tumor biopsy for PD-L1: Within 72 hours after the PD-L1 PET scanning, one or more tumour biopsies are obtained from metastatic lesion (-s). Core biopsy needles are used to increase chances to obtain representative material and to minimize the change for sample bias due to spatial heterogeneity in PD-L1 expression. The biopsy is performed by an experienced radiologist and will be image-guided with either ultrasound- or CT-directed modalities. The site of biopsy is determined by the radiologist and guided by the fused images obtained from the PD-L1 PET Investigation. Preferentially, material should be obtained from a lesion that shows PD-L1 positivity. In case of presence of both PD-L1 positive and PD-L1 negative lesions at the PD-L1 PET, when technically feasible and deemed safe by the radiologist, material is obtained from both a positive and a negative lesion. When available, material from the primary tumor is retrieved by the pathologist for review and to perform PD-L1 IHC.

The biopsied material is transported to the pathology department at the hospital where the biopsy is taken and managed according to clinical routine. Material from the primary tumor (when available) is collected. The Ventana SP142 antibody is used to assess PD-L1 status, where PD-L1 positivity is defined as positive PD-L1 expression on ≥1% of immune cells.

After performing IHC, whole slide PD-L1 stained tissue/core biopsy sections, including a positive and negative control, are scanned by 40x magnification (Hamamatsu or similar), and sent to the study pathologist for central revision.

Blood samples: Circulating PD-L1 mRNA: Exosomes are released from normal and from cancer cells, representing the phenotype of their originating cell by their content of RNA, DNA and proteins. PD-L1 gene expression from exosomally derived RNA was performed by Raimondi et al (16), demonstrating that high levels of PD-L1 mRNA expression at baseline were correlated with greater response to atezolizumab plus nab-paclitaxel, and that partial or complete response was positively associated with decrease in mRNA expression and improved overall survival. In our study, exosomal mRNA expression will be performed with the same strategy using the exoRNeasy kit (Qiagen) and Bio-Rad QX100 ddPCR.

Foundation One Liquid CDx: Material for Foundation One liquid biopsy is collected at baseline and at the first therapy evaluation, 8 weeks after treatment start. FoundationOne CDx™ (Foundation Medicine, Penzberg Germany; FoundationOne Liquid Cdx test) FoundationOne Liquid CDx is a next generation sequencing based in vitro diagnostic device that analyses 324 genes. Substitutions and insertion and deletion alterations (indels) are reported in 311 genes, copy number alterations (CNAs) are reported in 310 genes, and gene rearrangements are reported in 324 genes. The test also detects tumor fraction and the genomic signatures blood tumor mutational burden (bTMB) and microsatellite instability high (MSI-H) status. FoundationOne Liquid CDx utilizes circulating cell-free DNA (cfDNA) isolated from plasma derived from the anti-coagulated peripheral whole blood of cancer patients. For this sample, a separate informed consent form will be retrieved from the study participant.

Feces microbiome analyses: Feces samples are collected at baseline and at the first treatment evaluation after two 4-weekly treatment cycles. The aim of this project is to evaluate potential therapy-modifying factors verified from the gut microbiota. Several studies have shown that efficacy of ICI treatment is influenced by the microbiome, e.g. the gastro-intestinal/colonic bacterial flora (17, 18). Both pre-clinical as well as clinical data have shown that presence of certain forms of microbiome constitute as well as recent and current use of antibiotics are related to a lack of ICI treatment effect. Clinical data on antibiotic and proton pump inhibitor use will be monitored.

#### 3. Clinical Outcome Assessments

All patients will undergo a baseline CT of the chest-abdomen to be able to perform a treatment evaluation after 2 courses (8 weeks) with the systemic therapy as defined above. In addition, routine blood tests are performed, at the discretion of the treating physician. For the PD-L1+ patients who are treated with atezolizumab, additional blood tests aimed at measuring toxicity from this drug will be performed aimed to detect signs of immune-mediated toxicities. A CT/MRI of the brain will be performed in case of a clinical suspicion for CNS metastases, or when PD-L1 positive lesion(s) are noted on the baseline PD-L1 PET scan.

Toxicity management, including supportive medication and dose reductions, is done at the discretion of the treating physician in conjunction with the Principal Investigator and according to the recommendations in the study protocol.

### Outcomes

Objectives and endpoints are summarized in Table 2. The primary endpoint is the concordance between the current reference standard for PD-L1 testing (IHC on a tumor biopsy) and the experimental method (PD-L1 PET). Secondary endpoints are related to heterogeneity in PD-L1 receptor expression and the therapy-predictive value of PD-L1 PET. Exploratory endpoints relate to detection of metastases in sanctuary sites such as the central nervous system, relation with circulating biomarkers and the possible relation with uptake on PD-L1 PET and the development of immune-mediated toxicity.

**Table 2:** Objectives and endpoints of the MIMIR-mTNBC trial.

|  | Objectives | Endpoints |
| --- | --- | --- |
| *Primary* | Establish the statistical agreement (by means of Cohen's kappa coefficient) between PD-L1 status by the experimental method (PET/CT with <sup>89</sup>Zr-atezolizumab) and the current reference standard (IHC on a tumor biopsy or resection specimen). | Level of statistical agreement by means of Cohen's kappa coefficient between PD-L1 IHC (positive defined as expression ≥1% on immune cells with SP142) and PD-L1 PET/CT (PD-L1 positivity is defined as having at least one lesion with radiotracer uptake over the background uptake). |
| *Secondary* | Evaluate the therapy-predictive value of <sup>89</sup>Zr-atezolizumab PET/CT, compared to the current reference standard (PD-L1 IHC) for treatment with nab-paclitaxel, carboplatin + atezolizumab. Three patient groups are defined, and treatment outcomes (progression free survival, disease response rate, treatment discontinuation rates) will be compared:<br>1) IHC positive and PET positive,<br>2) IHC positive and PET negative,<br>3) IHC negative and PET positive. | Progression free survival, disease response rate and treatment discontinuation rates in three patient groups<br>IHC positive and PET positive,<br>IHC positive and PET negative,<br>IHC negative and PET positive. |
|  | Investigate inter- and intra-lesional differences in PD-L1 positivity by <sup>89</sup>Zr-atezolizumab radiotracer uptake | Discordance in <sup>89</sup>Zr-atezolizumab between different sites and within metastatic sites in the body. |
|  | Investigate the therapeutic role of atezolizumab for ER-low, PD-L1 positive breast cancer (ER expression 1-10%). | Progression free survival, disease response rate and treatment discontinuation rates in patients with ER-low mBC treated with nab-paclitaxel, carboplatin and atezolizumab. |
|  | Investigate the possible role of <sup>89</sup>Zr-atezolizumab PET/CT as a method of detecting otherwise (with regular CT-chest-abdomen) occult metastatic sites including sanctuary sites such as the central nervous system (CNS) | Percentage of <sup>89</sup>Zr-atezolizumab uptake in sites, not previously determined on the routine radiological investigation with CT, as a measure of cancer spread determined on whole body <sup>89</sup>Zr-atezolizumab PET/CT in and different metastases. |
| *Exploratory* | Explore mechanisms for possible discordance between PD-L1 positivity by IHC and <sup>89</sup>Zr-atezolizumab radiotracer uptake on PET/CT | Differences in immune infiltrate and tumor cells in biopsied sites with discordance between PD-L1 IHC and PD-L1 PET |
|  | Study the genetic profile of circulating (fractions of) tumour cells to predict treatment responses and monitor changes in biological behaviour by means of ctDNA assessed by Foundation One liquid biopsies CDx-analysis at baseline and at the first treatment evaluation, with a specific focus on identifying patients with an enriched 'Immunotherapy pathway' in relation to PD-L1 status on PET and IHC and levels of circulating PD-L1 mRNA measured in exosomes at baseline and the first treatment evaluation | Levels and composition of circulating biomarkers and relation to treatment response (PFS, disease response rate) and PD-L1 positivity by PET and IHC<br>- Levels of circulating PD-L1 mRNA<br>- Gene expression signals in liquid biopsies |
|  | Explore off-target effects of ICI by means of studying <sup>89</sup>Zr-atezolizumab radiotracer uptake in organs at risk for immune-mediated toxicities as well as studying the composition of and changes in the intestinal microbiome. | Rates (and severity according to CTC-AE) of immune-mediated side effects in relation to <sup>89</sup>Zr-atezolizumab tracer uptake in organs at risk for immune-mediated toxicities. |

### Data management

Data are registered using an electronic Case Report Form (eCRF). Monitoring is performed according to Good Clinical Practice (GCP) guidelines. The eCRF provides data on patient demographics, tumor biological and treatment characteristics, as well as and data on follow-up. Data are managed by the Clinical Trial Office at Centre for Clinical Cancer Studies, Karolinska University Hospital, Stockholm, Sweden. Recorded information is pseudonymised. The MIMIR-mTNBC trial is registered at www.clinicaltrials.gov (NCT05742269). Ethical permission has been obtained by the Swedish Ethical Review Authority and Medical Product Agency (Dnr 5.1.1-2022-73437). All participants must provide their written informed consent to participate in this trial.

### Follow-up

The trial ends for each participant followed for 30 days after disease progression but also for participants who die, withdraw consent, or are lost to follow-up.

### Sample size

This is an exploratory phase II non-randomized study with the primary aim to assess the statistical agreement (by Cohen’s kappa coefficient) between the current gold standard (PD-L1 IHC by SP142, where PD-L1 positivity is defined as expression ≥1% on immune cells) and the experimental method with PD-L1 PET (PD-L1 positivity is defined as having at least one lesion with radiotracer uptake over the background uptake). Approximately 40% of cases are expected to have PD-L1 positivity by means of the reference standard with PD-L1 IHC, based on the data from the IMpassion130 and Keynote 355 trials. We regard a kappa of 0.80 as having a very good statistical agreement; 64 patients are needed to be able to estimate the kappa with a precision of +/- 0.15.

### Statistical methods

Continuous data will be summarized using descriptive statistics, where the following parameters will be reported: number of evaluable and missing observations, mean and standard deviation, median, quartiles, and extreme values (minimum and maximum) unless stated otherwise. Categorical data will be presented as number and percentage of subjects. Tests will be two-sided and performed at the 5% significance level unless stated otherwise. All eligible patients will be included in the analysis of the primary, secondary and exploratory objectives. The safety analysis set will contain all subjects who have been exposed to the investigational products ^89^Zr-atezolizumab and atezolizumab.

### Independent safety analysis

An Independent Data Monitoring Committee will execute the following monitoring plan and can recommend termination of the study based on the pre-defined stopping criteria. A patient will be withdrawn immediately if she/he withdraws informed consent at her/his own request, unacceptable toxicity occurs, not manageable by symptomatic therapy, dose delay or dose modification, she/he does not comply with the instructions given by the study personnel, pregnancy occurs, the medication has to be discontinued for medical reasons, including severe side effects, other disease, need of other cancer treatment. Patients who have been withdrawn will continue to be tracked and assessed (unless the patient explicitly declines). Entire trial: The study can be stopped due to Adverse Events whose frequency, severity or gravity justify discontinuation, as determined by the Principal Investigators.

### Time plan

Recruitment will commence March 2023 at the Karolinska Comprehensive Cancer Centre, Sweden. Further participating sites will be added according to interest and need. Patient enrolment is estimated to be completed by the end of 2025, and first results on the primary endpoint are reported in 2026.

## Discussion

The current standard of care for treatment of metastatic or irresectable triple negative breast cancer is the combination of ICIs with a chemotherapy backbone for patients with a PD-L1 positive tumor biopsy with the SP142 (for atezolizumab) or 2CC3 IHC antibodies (pembrolizumab) (1-4), leading to an improved overall survival compared to chemotherapy alone.

There is a clear clinical need to improve the selection of patients for this treatment combination to optimize the risk-benefit balance at an individual and societal level, as several downsides exist with the currently used reference standard therapy-predictive biomarker of PD-L1 IHC. This relates amongst others to the need for invasive tumor biopsies which are burdensome for patients and sometimes practically not feasible, the discordance in PD-L1 status between (different sites of) metastases and primary tumors and the limited representation of the results of a tumor biopsy from a solitary disease localisation compared to the whole-body burden of disease.

PET-imaging with tracers representing the targets of treatment is a possible way to evade these limitations and enables a real-time assessment of the target expression in the whole body which can be repeated when needed.

^89^Zr-atezolizumab is the first radiolabelled PD-L1 PET-tracer that has been evaluated. Warranting results from the first in human study provide the rationale for the MIMIR-mTNBC trial, where we hypothesize that PD-L1 PET imaging with ^89^Zr-atezolizumab may be a more reliable and patient-friendly alternative to the current reference standard with PD-L1 IHC on a tumor biopsy. In addition, studying whole body expression of PD-L1 is thought to provide an enhanced insight in tumor biology and development of response to treatment as well as immune-mediated toxicity. This was very recently very elegantly shown in a clinical trial with a ^89^Zr-labeled CD8 tracer ^89^ZrED88082A, where a CD8-PET was done prior to and after 30 days of treatment with ICI (19). An enormous heterogeneity in CD8^+^ T cell distribution and pharmacodynamics within and between patients was noted, revealing and visualizing the effects of ICIs on T-cell recruitment throughout the body.

A few trials with ^89^Zr-atezolizumab have been initiated, according to a search at the website Clinicaltrials.gov (searched February 14^th^ 2023); NCT04006522 in patient with metastatic renal cell carcinoma, NCT03850028 in patients with malignant lymphomas and NCT05404048 in patients treated with CAR T-cell therapy. One imaging sub-study to a clinical trial in patients with metastatic lobular breast cancer with ^89^Zr-atezolizumab has been registered at clinicaltrials.gov (NCT04222426); the main study was terminated after including only one patient.

Several imaging trials with other radiolabelled PD-L1 and PD-1 antibodies are currently ongoing in different patient cohorts; ^89^Zr-pembrolizumab (active trials: NCT02760225, NCT03065764), ^89^Zr-durvalumab (NCT03610061, NCT03829007) and ^89^Zr-avelumab (NCT03514719).

Taken together, PET-imaging of PD-L1 and PD1 expression as well as other mediators in the response to ICIs may augment insights in the mechanisms of response to these drugs. In addition, this may provide a real-time whole-body assessment of the target of treatment and thereby be used as a therapy-predictive biomarker. The results of the MIMIR-mTNBC may add in the body of evidence of this area, with the prime focus to investigate whether the PD-L1 status by PET correlates to the PD-L1 status determined with IHC on a tumor biopsy, which is the current standard of care for patients with mTNBC.

In conclusion, in the MIMIR-mTNBC trial, we aim to compare the level of statistical agreement between the current reference standard to assess PD-L1, i.e., on a tumor biopsy by means of IHC, to the experimental method of PD-L1 PET imaging with ^89^Zr-atezolizumab. Our academic-initiated trial addresses a highly unmet medical need and may open new applications for superior and specific non-invasive tumor characterization. If ^89^Zr-atezolizumab PET/CT demonstrates a sensitivity and specificity rate at least equal to the currently used reference method (IHC) it should impact patient management by selecting optimal patients for treatment with ICIs. In addition, such a precision medicine tool will substantially benefit the health economy by reduce cost for expensive therapies in patients without expected benefit and only side effects.

## Data Availability

This is a manuscript presenting a study protocol without preliminary data and therefore, this is not applicable here.

## Acknowledgements

The study team is thankful for the scientific input from Professor E.G.E. de Vries from the University Medical Center in Groningen, the Netherlands and the material support from Roche. The administrative support of the Clinical Trials Office at the Karolinska University hospital, Stockholm Sweden is greatly acknowledged.

## Data Availability

All relevant data needed to replicate the trial can be found in the study protocol and through communication with the Principal Investigator.

